# Social behaviours and contact patterns across the 2020/21, 2021/22 and 2022/23 winter seasons in the UK, and associations with symptoms

**DOI:** 10.1101/2025.08.05.25333033

**Authors:** Elisabeth Dietz, Emma Pritchard, David W. Eyre, Tim Peto, Nicole Stoesser, Philippa Matthews, Tom Fowler, Conall Watson, Thomas House, Koen B. Pouwels, Ann Sarah Walker, the COVID-19 Infection Survey

## Abstract

The SARS-CoV-2 pandemic had a large impact on social mixing in the UK. This study analysed data from the Office for National Statistics Coronavirus Infection Survey to examine changes in contact patterns and self-reported symptoms through three winter seasons from 2020 to 2023. Using Generalised Additive Models, we estimated levels of various contacts over time, accounting for age, sex, ethnicity, and deprivation percentile, and compared these to trends in self-reported symptoms. Our estimates indicated steady increases in physical contacts from quarter-4 2020 to the end of the study in quarter-1 2023, with notable variation in age-specific trends. School closures and holiday periods had substantial impacts on contact patterns, particularly for children. Prevalence of reported symptoms also increased steadily over time, but varied much more within-season than most contacts; specifically, the relative increase in respiratory symptom prevalence during winter peaks between seasons was much larger than increases in contacts. Our estimates also suggested that while age played a crucial role in both contact patterns and symptom reporting, the effects of deprivation were less clear and far smaller. Our findings provide insights into changes in behaviours and symptoms during the pandemic, which may help inform future public health policy and infection modelling.

## Introduction

The SARS-CoV-2 pandemic had a large impact on the contact patterns of individuals in the UK. Public health policies and guidance, including on social distancing measures, along with personal choices, affected the nature and frequency of interpersonal contacts. Understanding how social behaviour changed over time during the pandemic is important to both public health policy and infection modelling in the future. Various studies have attempted to estimate such changes, including the CoMix study in the UK, highlighting substantial reductions in contacts early in the pandemic compared to pre-pandemic levels^[1]^. However, while a later snapshot study was conducted between November and December 2022, CoMix data collection generally ended in spring 2022^[2]^, and less is generally known about contact behaviours during the later stages of the pandemic.

Using digital contact tracing data from the NHS COVID-19 app, Kendall et al.^[3]^ (2024) estimated contact rates along with the reproduction number of SARS-CoV-2, for a longer time period including early 2023. However, while this study provided estimates for the later period of the pandemic, contact rates were estimated for SARS-CoV-2-positive index cases, which may exhibit higher contact rates than the general population. Data was also not available for children, given the apps’ restricted availability to those 16 years and older. The lack of demographic data also means that evaluating generalisability of results is challenging, and that the influence of factors such as age on social mixing could not be estimated. Quantifying age effects, and estimating trends for children, adolescents, and younger and older adults, may prove valuable for future infection modelling.

The Office of National Statistics (ONS) Coronavirus Infection Survey (CIS) provides an opportunity to study longer-term changes in contact patterns throughout the pandemic in the UK, including the effect of demographic variables like age, based on a broadly representative sample. This large longitudinal household survey collected data across three UK winter seasons between 2020 and 2023, facilitating comparisons of contact levels across varying SARS-CoV-2 prevalence and official social distancing guidance. Here we therefore estimated levels of a range of contact-related variables over time and compared them to levels of seven key symptoms.

## Methods

CIS was a large longitudinal household survey, covering a sample broadly representative of the wider UK population^[4-6]^. Households were randomly approached to participate, primarily from address lists. For everyone aged 2 years or older in the household, an approach was made for relevant written consent to have study assessments (including questionnaires covering demographics, symptoms, and contacts^[7]^), and provide self-taken nose and throat swabs for SARS-CoV-2 polymerase chain reaction (PCR) tests^[8]^, at approximately monthly intervals. Parents/carers provided consent for those aged under 16 years, and those aged 12-16 also provided written assent. Assessments were conducted by physical home visits by study workers until July 31^st^ 2022, and remotely, either online or by telephone, with swab kits posted to participants and returned by post/courier, from July 10^th^ 2022 onwards (following a small pilot starting May 2022). The study received ethical approval from the South Central Berkshire B Research Ethics Committee (20/SC/0195).

This analysis included study assessments between August 1^st^ 2020 and March 5^th^ 2023 (end of the study), where the SARS-CoV-2 PCR test result was negative (95.4% of all assessments during this period) and data for the seven symptoms studied throughout the survey was complete (99.5% of all assessments during this period). SARS-CoV-2 positivity was mostly <2% throughout this period, except for short periods during Omicron waves^[9]^. Other excluded study assessments either had void (2.2%) or invalid (0.5%) test results. Analysis was restricted to SARS-CoV-2-negatives to avoid influence from test positivity on contact patterns (e.g. due mandatory isolation) and symptoms. Observations before August 2020 were excluded as information on participant contacts was only solicited from late July 2020.

### Social behaviours and contacts

At every assessment, participants were asked how many non-household (“not living in your own home”) contacts they had in the last seven days. Responses were as categories (0, 1-5, 6-10, 11-20, or 21 or more contacts), separately based on the contacts’ ages (children and young adults <18y, adults 18-69y, and older adults ≥70y) and nature (physical contacts and socially distanced contacts). Physical contacts, which were the focus of our analysis, were defined by direct in-person contact (including while wearing personal protective equipment), specified as “e.g. a handshake, hug, or personal care”. In contrast, socially distanced contacts were defined by direct in-person non-physical contact, i.e. interaction while maintaining social distancing. Socially distanced contacts were on average higher than physical contacts by ∼4-6 weekly adult contacts, ∼2-3 weekly child contacts, and ∼1-2 weekly older adult contacts, but associations with factors were similar so detailed analyses are not shown.

Participants were also asked, on average over the last 7 days, how many hours (0-24) they spent within 2 meters of someone else in their home (including sleeping). They were also asked how many times the following had occurred in the last 7 days (0, 1, 2, 3, 4, 5, 6, or 7 or more times): receiving a visit from a non-household contact inside their home, visiting inside someone else’s home, going outside home for shopping, and going outside for socialising (including visiting restaurants). Participants were also asked about using face coverings, with separate questions for work/education and enclosed public spaces (i.e. shops or public transport). Questions asked whether they “generally wear any kind of face covering or mask” when at these locations, “because of COVID-19”, with responses recorded as “Not going [to these locations]”, “Yes, always”, “Yes, sometimes”, “Never”, and “My face is already covered for other reasons (e.g. religious or cultural reasons)”. “My face is already covered” was grouped with “Yes, always”. We analysed the percentage reporting always using face covering (“Yes, always”) and using face covering sometimes at minimum (“Yes, always” and “Yes, sometimes” combined), with those reporting not going to the location in question excluded.

Due to differing starting points for data collection, these contact and social behaviour variables were analysed from August 1^st^ 2020 (physical contacts), January 1^st^ 2021 (hours within 2m of someone else, number of times received visitor/visited someone else), January 15^th^ 2021 (face covering), and June 1^st^ 2021 (number of times outside for shopping/socialising) respectively.

### Symptoms

At each assessment, participants were asked whether they had experienced specific symptoms during the last seven days. 12 different symptoms were solicited initially, with a further 4 and 7 added in September 2021 and January 2022 respectively. Our analysis included 7 symptoms solicited from the survey start (cough, sore throat, shortness of breath, fever, headache, muscle ache (myalgia), and weakness/tiredness (fatigue)), which are included in the European Centre for Disease Prevention and Control definition of influenza-like illness^[10]^ (co-presence of ≥1 respiratory symptom (cough, sore throat, shortness of breath), and ≥1 systemic symptom (fever, fatigue, headache, myalgia)).

### Statistical methods

All outcomes (each contact/behaviour and symptom variable) were modelled with Generalised Additive Models (GAMs), using the R *mgcv* package^[11]^. All models included smooths representing interactions between age and calendar time, and between deprivation percentile and calendar time (i.e. allowing the associations between age and each outcome, and deprivation and each outcome, to change over time). These tensor products were modelled with thin plate splines and k=30 basis functions^[12]^. The number of basis functions was selected to keep average daily predictions within 0.25 contacts of those estimated from a model with k=40 (approximately half the age range included in the models), balancing effective degrees of freedom and computational constraints given the large sample size^[13]^.

Participant age was recorded in years at each assessment (truncated in models to 5-85 due to low numbers outside this range). Deprivation percentile was derived separately for each country, based on participant residence postcode and the respective indices for multiple deprivation (IMD)^[14-17]^. Fixed effects for sex (male/female), country of residence (England/Scotland/Wales/Northern Ireland), ethnicity (collapsed to white/non-white due to low numbers) and day of the week were also included in all models. Although the questionnaire solicited responses about symptoms and contacts in the past seven days, we found reporting behaviour varied significantly by study assessment day, prompting the inclusion of fixed effects for day of the week. Models were estimated separately for each outcome and season (2020/2021, 2021/2022, and 2022/2023) defined by the UK financial year (April-March) to avoid splitting the winter season between calendar years.

The percentage reporting each of the seven symptoms was estimated using negative binomial models with a log link (to allow for over-dispersion), as was the percentage using face covering (separate models for each location and for “Always” and “Always or Sometimes”). The number of hours spent within 2 meters of someone else was estimated using linear regression. The mean number of physical contacts (with those aged <18y, 18-64y, 75y+) was estimated using interval censored regression (“cnorm” family in *mgcv*), given that response categories were interval or right censored. Variables from questions asking “the number of times …” were also estimated using interval regression, given that responses were right censored for those reporting “7 times or more”.

To account for the effects of two specific changes to survey delivery on symptom reporting, age-specific offsets were included in the symptom models. The first was a change in survey layout on December 14^th^ 2020; the second was data collection transitioning from study workers to online/telephone during July 2022. Both changes generally increased symptom reporting, with larger increases at older ages for several symptoms **(Supplementary Figure S1)**, and hence offsets to adjust for these changes were included in all symptom models, so that estimates reflect the levels that would have been reported with an online/telephone survey (details in **Supplementary Methods**). Offsets were not included in models for contact variables due to the lack of evidence of substantive effects of these survey changes on contact reporting behaviour **(Supplementary Figure S2)**.

## Results

10,737,078 assessments from 532,802 unique participants were included in analyses (median 22 assessments per participant, IQR 15-26). The median participant age across assessments was 51 years (IQR 32-66 years) and the median deprivation percentile was 63 (IQR 39-83, higher=less deprivation), suggesting that the sample was somewhat older^[18]^ and less deprived than the overall UK population (both factors adjusted for in models).

### Physical contacts outside home involving adults

Trends in the number of reported physical contacts outside of the home varied substantially over time and age, with notable increases observed from the start of study period in the third quarter of 2020 (Q3-2020) to the end in Q1-2023 **(F1, Supplementary Figure S3)**. Young children and young adults were estimated to have the highest levels of physical contact with adults aged 18-69 years **(F2)**. At age 25, participants were estimated to report ∼4 weekly adult contacts in September 2020, increasing to ∼7 two years later. At ages 45 and 70, participants had fewer adult contacts, but these were also estimated to increase over the same time period, with around 2 additional contacts reported in September 2022 compared to September 2020, on average. Young children saw larger estimated increases in adult contacts over the same time period than older children, increasing from ∼3 to ∼6 at age 5 compared to from ∼2 to ∼4 at age 15. By spring 2023, the number of weekly adult contacts appeared to have stabilized, with higher levels reported at ages 5 and 25 (∼7-8 and ∼6-8 respectively) followed by age 45 (∼5-6) and ages 15 and 70 (∼4-5) **(F1)**.

**Figure 1.**
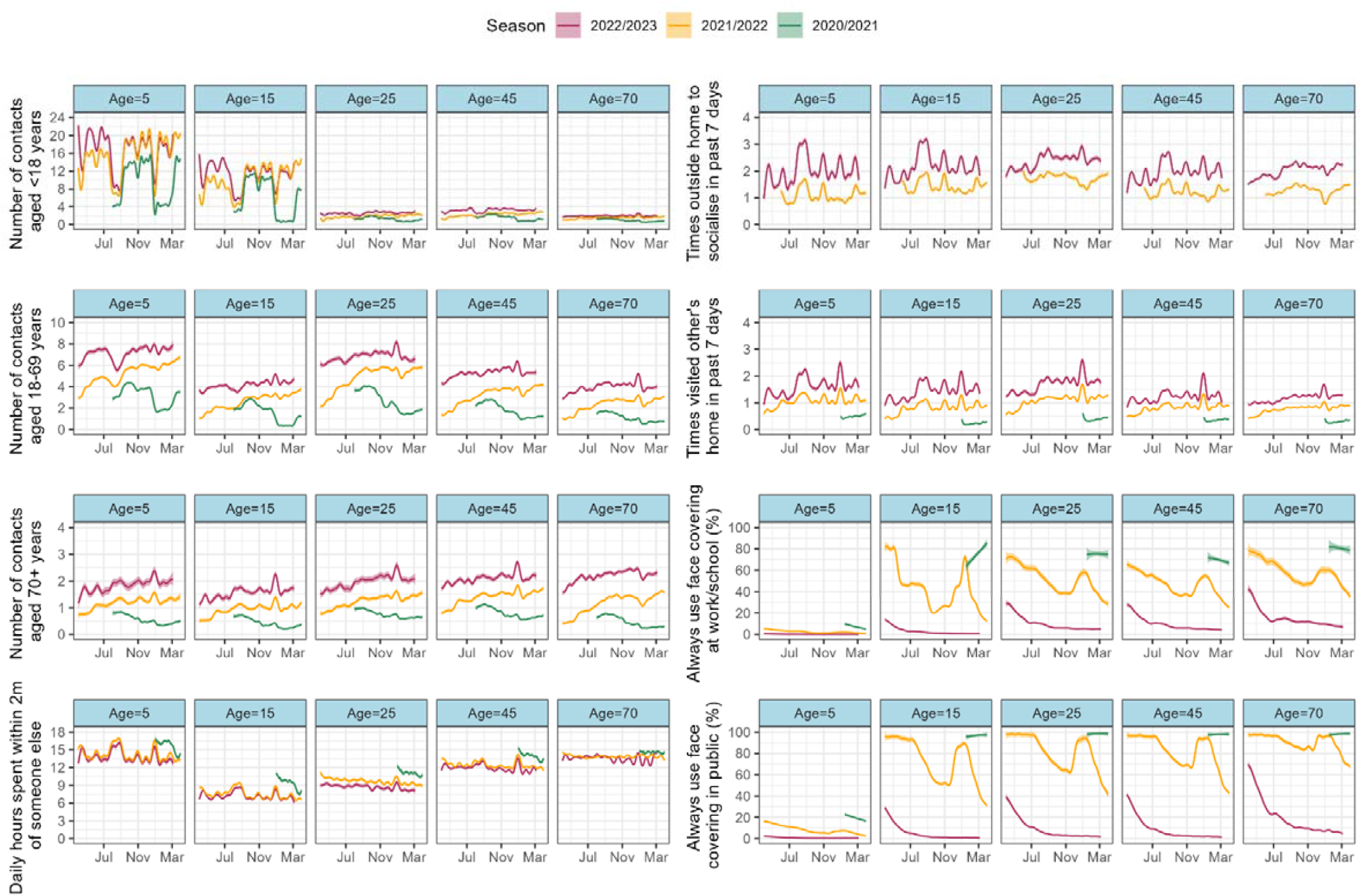
Trends in contact variables August 2020 – March 2023, by season at specific participant ages. NOTE: Estimates obtained at the median/mode in-sample levels of the fixed effects (deprivation percentile=63, sex=female, ethnicity=white, country=England, DOW=Wednesday). Estimates for the remaining contact variables are shown in **FS3**. Fixed effect estimates are shown in **FS4**.

Levels of physical contact outside of the home with older adults aged 70+ were generally lower, but also increased over the same time period **(F1)**. Across ages, around one additional older adult contact was reported in September 2022 compared to September 2020, with slightly larger increases estimated for participants aged 70. A “Christmas effect” of temporarily increased contact levels was clear in the two last seasons (22/23 and 21/22 (emergence of the Omicron BA.1 variant)) for both adult and older adult contacts across most participant ages. In contrast, December 2020 saw a sharp drop in contact levels, particularly visible in adult contacts for child participants. This time period coincides with the emergence of the Alpha variant, which was followed by the introduction of tiered restrictions and a national lockdown (including school closures) in England from early January.

At age 5, children were estimated to have dropped from ∼4 weekly adult contacts in autumn 2020 to ∼2 between January and March 2021. This contrasts with levels of around ∼6 and ∼7 over the same months in the two later seasons. For older children aged 15, adult contacts (outside the home) were also estimated to drop from ∼2 to almost zero over this time period.

### Physical contacts outside home involving children

Levels of physical contact outside of the home between children were similarly at very low levels between late December 2020 and March 2021 **(F1)**. Generally, physical contact with children aged below 18 was highly age-dependent, with fluctuations corresponding approximately to school term for school-aged children. At ages 5 and 15, term-time peaks contrasted with troughs during summer and Christmas holidays. Contact levels also reduced noticeably during the shorter vacations (autumn half-term, spring half-term, Easter holiday and late spring half-term), but only by approximately half of the amount of the longer vacations. At ages 5 and 15, autumn term peaks in physical contact with other children/adolescents under 18y were higher in 21/22 and 22/23 seasons than in the first 20/21 season, when schools were re-opening after the first pandemic-related closures (March-July 2020, before this study started). As above, the spring term peaks visible in later seasons did not occur in the 20/21 season, indicating the impact of the second period of school closures. During 2021, summer term-time peaks were still somewhat reduced, appearing to stabilise from autumn 2021 onwards. Physical contacts between adult participants and children were generally at lower levels (<4 weekly child contacts at age 45 compared to >12 term-time weekly child contacts at age 15 during the latest season), but gradually increased from the start of the study period. A drop in non-household child contacts during the school-closure period December 2020–March 2021 was the most visible at age 45, highlighting potential associations between parents and children.

### Physical contacts within households, recreational activities, and face coverings

The number of hours within 2 meters of someone else in the home appeared to follow an inverse trend to physical contacts with other children (and thus term dates) at age 5 **(F1)**. For older participants, the number of reported hours was broadly stable over time, with visible “Christmas effects” and marginal decreases from 21/22 to 22/23. Notably higher levels were observed between December 2020 and March 2021 across ages, suggesting an impact of tiered restrictions on socialising outside the home resulting in greater time spent with others within the home. At ages 5 and 10, the number of times individuals reported socialising outside home was estimated to follow a similar pattern, with clear summer holiday, and smaller half-term, peaks. As above, those aged 45 appeared to have somewhat similar trends to those seen for children. Across ages, the number of times people socialised outside the home was higher in 22/23 than 21/22, with an increase of around ∼1 additional trip per week. Trends were relatively similar for trips outside home for shopping, receipt of visitors at home **(Supplementary Figure S3)** and visits to others’ homes **(F1)**. Shopping trips were more frequent in older than younger participants, and “Christmas effects” were visible in visiting trends across ages **(F1, F2, Supplementary Figure S3)**. The use of face covering at work/school and in enclosed public places generally also decreased from Q1-2021 to Q1-2023, with a temporary peak between November 2021 and March 2022 (emergence of the Omicron BA.1 variant **(F1)**. The use of face covering was somewhat higher in enclosed public places than at work/school, and lower in children. (**F1)**.

**Figure 2.**
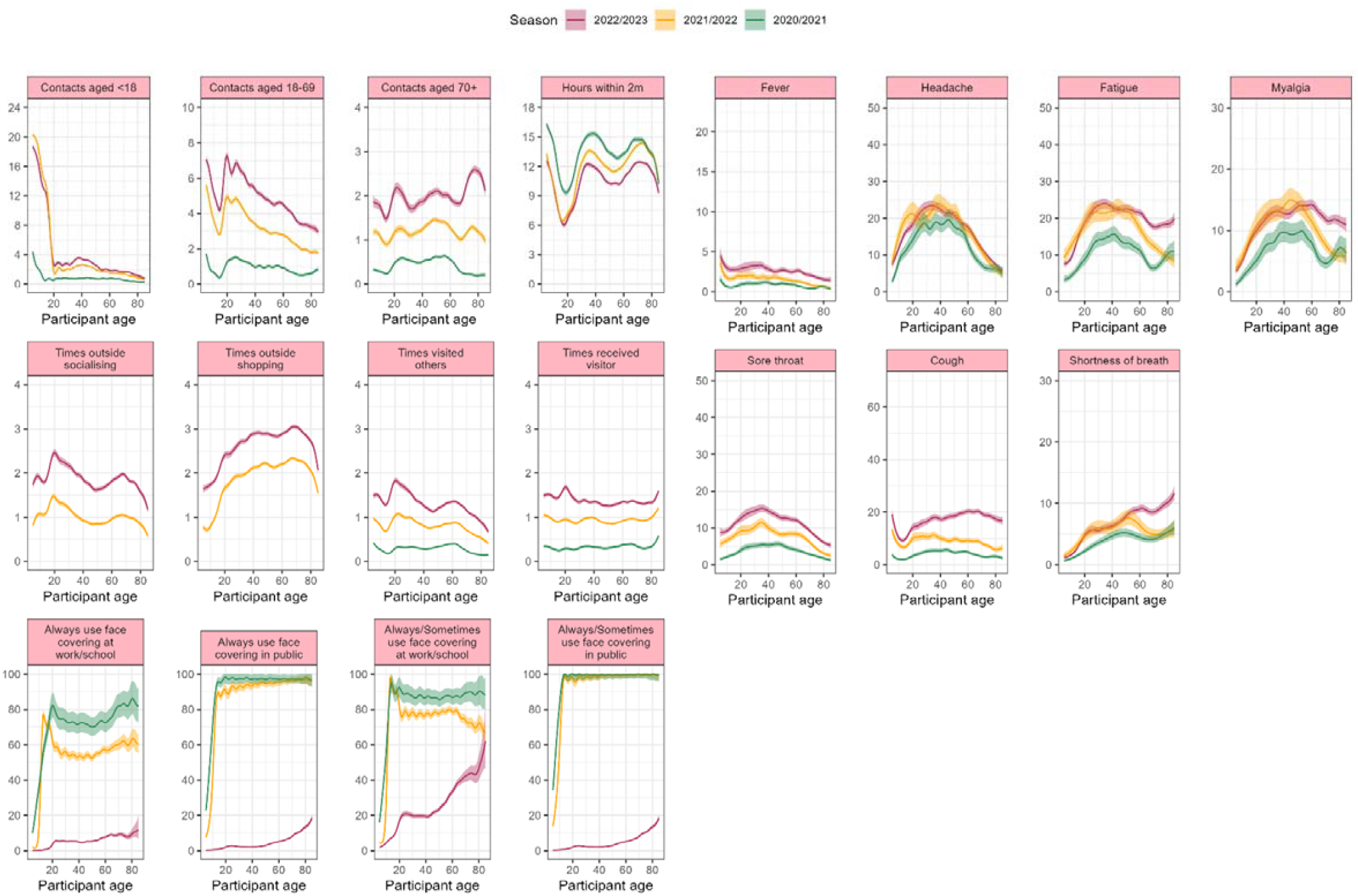
Effect of age on various contact variables (left) and symptoms (right), by season. NOTE: Estimates obtained at the median/mode in-sample levels of the fixed effects (deprivation percentile=63, sex=female, ethnicity=white, country=England, DOW=Wednesday) and January 15^th^ (2021, 2022 and 2023). Fixed effect estimates are shown in **FS4**.

### Trends in self-reported symptoms

Levels of the seven self-reported symptoms included in this analysis (from the ECDC definition of influenza-like illness) also generally increased throughout the study period **(F3)**, even after adjusting levels upwards to account for changes to data collection **(Methods, Supplementary Methods)**. In particular, the “winter peak” observed across ages in December each year was much higher in 2022 than 2021 and 2020 for fever, sore throat, and cough. Headache, fatigue, myalgia and shortness of breath also had notable “winter peaks”, yet trends were broadly much more similar across the last two seasons studied. Generally, prevalence of self-reported symptoms varied considerably more over time than most contact variables, with clear peaks and throughs observed also for adult and older adult participants. The relative increase in respiratory symptoms during winter peaks being larger than increases to social behaviours may be explained by the relationship between contact rates and transmission. If the reproduction number (secondary cases per primary) scales linearly with contact patterns, transmission of respiratory illness will increase more than linearly as cases beget more cases.

**Figure 3.**
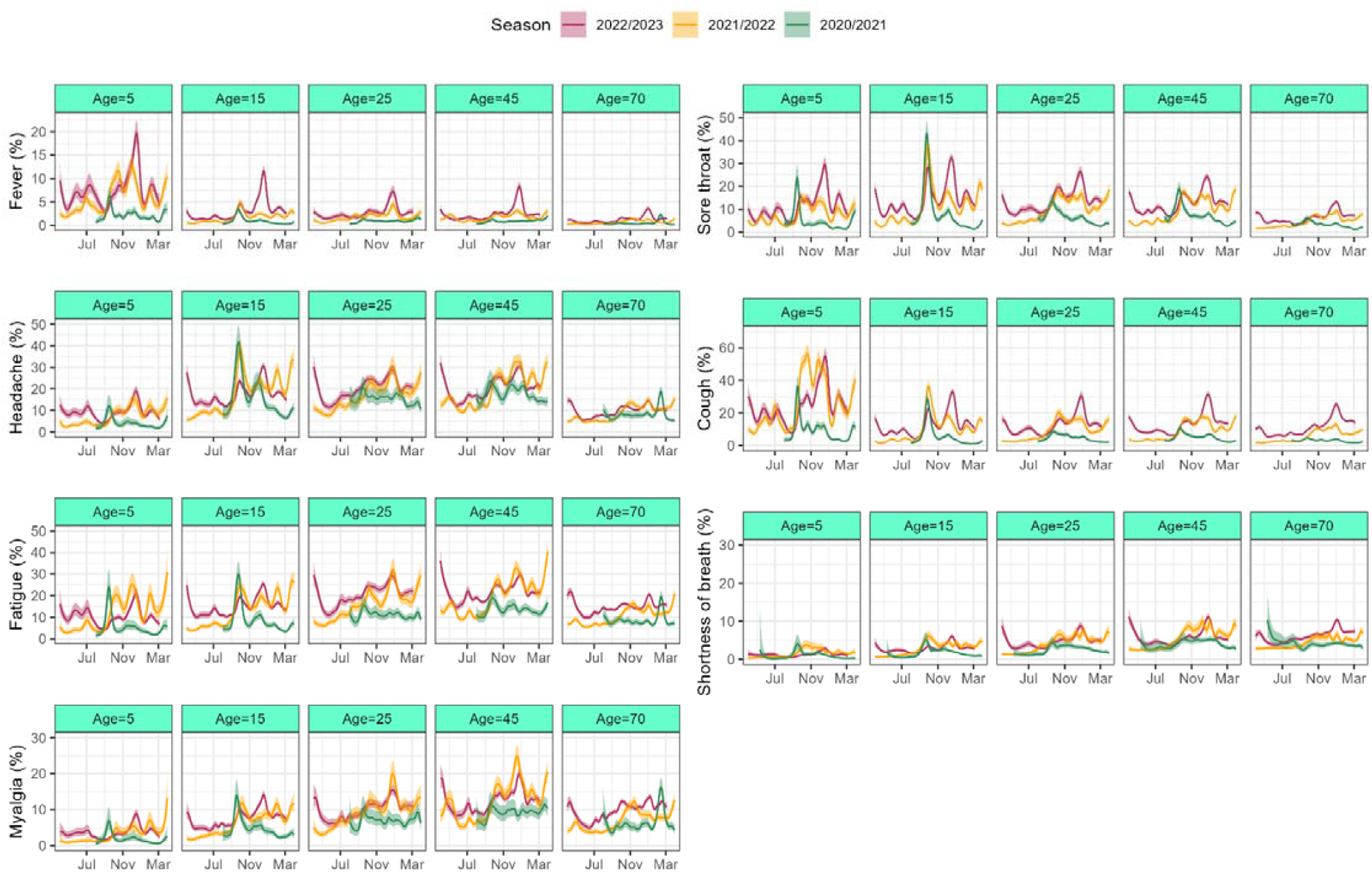
Trends in symptoms August 2020 – March 2023, by season at specific participant ages, adjusted for survey changes. NOTE: Estimates obtained at the median/mode in-sample levels of the fixed effects (deprivation percentile=63, sex=female, ethnicity=white, country=England, DOW=Wednesday). Fixed effect estimates are shown in **FS5**. Models for shortness of breath and myalgia were fit in 1-4 sections per season due to computational constraints, and merged using gradual smoothing over 2-week overlapping periods.

As for physical contacts, fluctuations in symptoms appeared influenced by school term dates, with clear peaks occurring after returns to school after holidays. At the beginning of the school year in October, there were general spikes in symptoms, particularly for children but also (to a smaller extent) for adults. Interestingly, this effect was particularly strong in the first 20/21 season for sore throat, headache, fatigue and myalgia, especially at age 15. Between January and March 2021, symptom reporting was generally lower across ages, with the spring half-term peak in mid/late February not occurring in this season as observed in later years. This reduction coincided with a reduction in contacts, as discussed above, during a period of tiered restrictions and lockdown following emergence of the Alpha variant. However, a small peak in systemic symptoms (fever, headache, fatigue and myalgia) was estimated around mid/late February for participants aged 70, which was not visible for other symptoms and age groups. This increase in systemic symptoms in early 2021 amongst older participants plausibly reflected short-lived side effects from COVID-19 vaccination which were tightly delivered according to age, and to the oldest participants during this time period.

### Impact of age on social behaviours

The relative effect of age on contact variables was very similar across seasons, although absolute levels varied **(F2)**. As above, the number of physical contacts with those aged below 18 years was much higher for children than adults, but with a small peak observed in adults aged around 40. Estimated physical contact levels with children were notably lower for those aged 5-10 years in the first season, likely due to effects being estimated on January 15^th^ when schools were closed (although children of keyworkers could attend). The number of physical adult contacts were high for very young children, but low for older children, then peaking around age 20-30 before decreasing for older ages. Participants generally had fewer contacts aged 70+, with a peak between participant ages 60-80 highlighting within-age-group contact. Interestingly, this peak was not visible for the 20/21 season, perhaps indicating the effects of the early pandemic on socialising for older individuals. Across seasons, young children reported a high number of hours spent within 2 meters of someone else, dropping substantially in adolescence, before rising again in young adults. Adults generally reported a high number of hours, potentially indicating both co-habitation and greater time spent at home. In general, participants were slightly more likely to report that they received a visitor than that they visited someone else, particularly for ages 60 and above. Young people aged around 20 had the highest rates of socialising outside the home, with another peak around ages 60-70. Shopping rates were generally higher in older participants with a peak around ages 40-70. Both shopping and socialising rates declined rapidly above age 70. Amongst older children and adults, the use of face covering at work/school or in enclosed public spaces did not vary much by age in the first two seasons. In the 22/23 season, rates were generally lower, but there was a clear increase with age.

### Impact of age on self-reported symptoms

Age effects on fever and cough were similar to those on adult contacts: higher in very young children, low in older children, and increasing again for younger adults **(F2)**. However, in the 22/23 season, rates of cough were also high for older adults, with a peak around 60 years. Headache and sore throat had similar distributions across age, with a clear peak around age 40, and lower rates in children and older adults. Age effects were also similar for fatigue and myalgia, with the exception of increased reporting amongst those aged 70 and above in the first season (estimated on January 15^th^ 2021), possibly related to short-lived vaccination effects. Rates of self-reported shortness of breath generally increased with age across seasons. In general, the relative effect of age on symptom reporting did not vary greatly between seasons, suggesting some stability. Although headache, fatigue, myalgia and sore throat peaked around age 40, we did not see this age group having notably distinct social behaviours. High rates of fever and cough both in children and middle-aged participants did however somewhat mirror trends in adult and child contacts, and it’s worth noting that those aged ∼40 may be likely to have young children, potentially influencing symptom rates **(F2)**.

### Impact of deprivation on social behaviours and self-reported symptoms

In general, the relative effect of deprivation percentile on social behaviours was unclear and of much smaller magnitude **(F4)**. Across the three seasons studied, those living in less deprived areas appeared slightly more likely to spend time at home within 2 meters of someone else, perhaps suggesting higher rates of working from home in this group, particularly in the first (20/21) season. There were also indications that those living in less deprived areas left home more frequently to socialise (including visiting restaurants), potentially associated with disposable income. The relative effect of deprivation on self-reported symptom levels was similarly not clear, with relative stability observed across deprivation percentiles and seasons for sore throat and cough in particular. However, there were indications that fever, headache, fatigue, myalgia and shortness of breath were more commonly reported by those living in more deprived areas. On January 15^th^ 2023, those living in the most deprived areas were estimated to have rates of fatigue 30.7% higher than participants living in the least deprived areas (absolute rates 26.4% vs. 20.2%).

**Figure 4.**
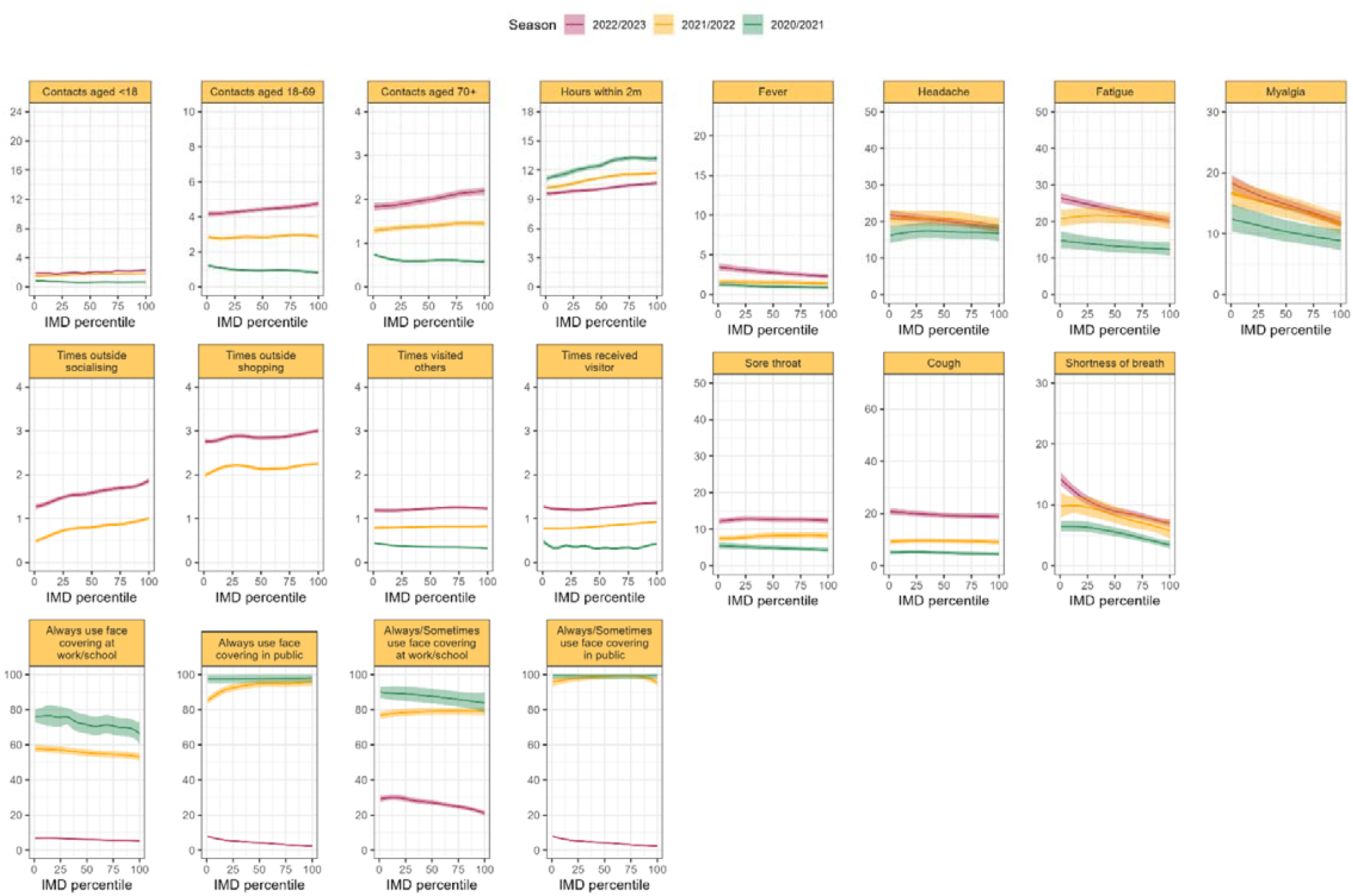
Effect of deprivation percentile on various contact variables (left) and symptoms (right), by season. NOTE: Estimates obtained at the median/mode in-sample levels of the fixed effects (participant age=53, sex=female, ethnicity=white, country=England, DOW=Wednesday) and January 15^th^ (2021, 2022 and 2023). Fixed effect estimates are shown in **FS4** and **FS5**.

## Discussion

Our analysis of CIS assessments over three winter seasons between 2020 and 2023 revealed multiple changes in contact patterns and symptom reporting. Importantly, the number of physical contacts participants reported weekly increased steadily through the seasons, along with other social behaviours such as receiving visitors and leaving home to socialise. Physical contact between children was much more common than other types of contact, with large peaks during term-time. Effects of widespread school closures from December 2020 to March 2021 could be seen in trends in physical contacts as well as symptoms, particularly for children. As expected, an increase in the number of hours spent within 2 meters of someone else was also evident during this time period, and although data was more limited on visiting behaviours at this time, there were indications of similar impacts from social distancing measures.

Apart from December 2020 to March 2021, and “Christmas peaks” in the two later seasons, physical contact with adults was relatively stable within-season, gradually increasing from August 2020 to March 2023. For participants aged 25, this amounted to an estimated increase from ∼4 to ∼7 weekly adult contacts, with a trough of <2 adult contacts early in 2021. Similarly, at age 45 participants were estimated to increase from ∼3 weekly adult contacts in August 2020 to ∼5 in March 2023. Except for physical contact between children, social behaviours did not exhibit large differences in time-trends across age, although absolute levels varied. For instance, older children reported the fewest adult contacts, compared to high levels in young children and young adults. The number of contacts aged 70+ was predictably higher for older participants aged 70 and above, although there were indications that within-age-group socialising was limited above the age of 70 in the first (2020/2021) season.

The final season studied differed from the two previous seasons in the large winter peaks observed for fever, sore throat and cough. This large increase in respiratory symptoms in winter 22/23 was particularly prominent in children, yet was not as visible for cough in the youngest children, who also saw high rates in winter 21/22. There were noticeable “back-to-school” effects around October in the reporting of fever, sore throat, headache, cough, fatigue and myalgia amongst children. Interestingly, such peaks could also be seen in sore throat, cough, headache, and fatigue amongst participants aged 45, appearing more prominent than amongst those aged 25 or 70. This observation highlights potential associations in transmission between school-aged children and their parents.

Young adults reported the lowest number of hours spent within 2 meters of someone else, and the highest rates of socialising, visiting others, and receiving visitors – along with high numbers of adult contacts. However, this age group did not report particularly high levels of any symptoms, perhaps except headache. In general, several symptoms were characterised by high levels amongst middle-aged participants. Headache, fatigue, myalgia, and sore throat were all estimated to be the most common amongst participants aged ∼40, with lower rates in children and young adults, as well as older adults. Nevertheless, there was a notable exception observed for fatigue and myalgia in the first season (20/21), where rates increased above the age of 70. This effect may be attributed to short-lived vaccination effects, which also appear to have led to temporary increases in fever and headache amongst older participants.

Fever and cough differed from the other symptoms by having high levels in young children below the age of 10. However, it is worth noting that these symptoms (signs) may be more easily identified by parents/carers filling out the questionnaire for children than more subjective symptoms such as fatigue. It is therefore possible that other symptoms are in fact under-reported in young children. Nevertheless, prior studies have found that parents may overestimate the severity of their child’s illness, suggesting that parent-reported symptoms could also overestimate true prevalence^[19]^. A systematic review by Smith et al. (2020) highlighted the role of parent and child affect, as well as parental expectations, in influencing symptom reporting, emphasising the need for additional research on symptoms in children^[20]^.

Overall, as expected, age played an important role in both symptom reporting and social behaviours: the key contribution of this study is detailed estimates of rates over time and by age that could be used to parameterise future infectious disease models. Interestingly, similar effects could not be identified for deprivation percentile, where effects were unclear or small in size. It has been suggested that deprivation could increase the likelihood of respiratory infections through increased exposure associated with higher likelihood of work outside the home in higher-contact professions, particularly in the context of SARS-CoV-2^[21,22]^. A prior study in England and Wales found that people living in more deprived areas were more likely to undertake essential activities such as using public transport and working outside the home, during autumn/winter 2021^[23]^. Our estimates also suggested a slight decrease with deprivation in the hours spent within 2 meters of someone else, perhaps suggesting lower rates of home working amongst those living in more deprived areas. However, there were no clear effects of deprivation on physical contact rates. If anything, participants living in less deprived areas reported slightly more adult contacts in the latest reason than those living in more deprived areas. Hoskins et al. (2023) similarly observed higher rates of non-essential social activities amongst those living in the least deprived areas as defined by IMD^[23]^. The estimated effects of deprivation on symptom reporting were also minor in comparison to those of age. There were however indications of higher rates of some symptoms amongst those living in more deprived areas, particularly in the latest season studied, and especially so for fatigue, myalgia, and shortness of breath. This finding is in line with prior work on the social determinants of health^[24-26]^.

A limitation to this study is that the definition of physical contact was constrained to “direct” contact with other people. People working in customer-facing jobs that could in fact involve increased risk of infection transmission (i.e. retail and grocery store workers, public transport staff, receptionists, and restaurant/café staff) may therefore not report high contact numbers, compared to in studies using ‘face-to-face’ definitions of contact^[21]^. Furthermore, the categories used to collect contact information were relatively wide, meaning that they may be insensitive to smaller changes. However, our use of interval regression for contact level estimation tried to account for this. Further, although absolute levels were somewhat higher, we observed similar trends in socially distant contacts. More generally, physical contact reporting may also be influenced by recall bias. While we were able to control for day-of-the-week effects (with higher contact numbers reported on Sunday and Monday compared to other days), it is possible that people may be less likely to remember all of their contacts at times when contact levels are higher. Although future studies could benefit from collecting more detailed contact data, this requires substantially more time commitment from participants; CIS prioritised scale as its primary objective was to estimate SARS-CoV-2 positivity.

Another key limitation is that area-based definitions of deprivation, as used in this study, may not always reflect individual or household levels of deprivation. The use of participant postcodes to map to relatively small areas with average populations between 700-2,100 is consistent with other studies^[14-17]^. However, it is possible that individual-level measures such as household income (not collected in CIS) could better identify suggested associations between deprivation, contacts, and symptoms. People working essential (potentially high-contact) jobs may also be less likely to take part in surveys due to time constraints. Another important limitation is the underrepresentation in this sample of specific demographics^[5,27]^, including younger children and non-white ethnicities, although our models attempted to adjust for these factors. It is also possible that both social behaviours and infection rates had not yet stabilised post-pandemic by the end of the 2022/2023 season, and that more recent data could shed further light on the associations between social behaviours, transmission, and symptom reporting. Effects may also have been obfuscated by the variation in social distancing measures across regions (models only controlled for country) over time. Another consideration is that mixing patterns may vary seasonally in ways that have not been captured by the survey, such as indoor vs. outdoor mixing and differences in ventilation (open vs. closed windows). Social desirability bias could also have influenced survey responses, particularly at times when restrictions on contacts were in place.

Lastly, it is possible that our adjustment for the two survey changes did not fully capture their effects on symptom reporting. Why effects of these survey changes differed across participant ages, as well as between symptoms, is not clear. Given the timing of the shift from study worker to digital data collection in July 2022, it is possible that effects from school vacation were almost completely confounded with the adjustment coefficients, despite our inclusion of time-trends. However, the estimated adjustments were smaller for school-aged children than older ages (40+ in particular) suggesting this may not have had a strong effect. Contact variables were not adjusted given the lack of evidence of strong effects from survey changes. Although models suggested that reported physical contacts between children decreased during the shift from study worker to digital data collection, the direction and timing of this effect is consistent with it being related to school vacations starting rather than reporting. Similarly, the estimated increase in the relative percentage of participants below age 20 sometimes using face covering at work/school after the transition to digital data collection can likely be explained by shifts in the denominator (fewer children going to school). Why participants aged 40 and above reported more hours spent within 2 meters of someone else after the transition to digital data collection remains unclear, but could potentially be associated with lower average working hours during the school summer holidays. Nevertheless, any effects on social behaviours were small in magnitude in comparison to effects on symptom reporting. The presence of such effects for symptoms, along with relatively consistent day-of-the-week effects for contacts, highlights how ignoring factors affecting reporting behaviour could bias estimates in long-term surveys such as CIS.

In conclusion, our analysis of the ONS CIS reveals significant changes in social contact patterns and symptom reporting in the UK over time during the first three years of the COVID-19 pandemic. Physical contacts increased steadily from 2020 to 2023, with notable age-specific variations and clear impacts from school closures and holiday periods. Symptom rates also increased, with winter peaks becoming more pronounced each year, particularly for respiratory symptoms. While age appeared to have a large influence on both social behaviours and symptom reporting, the association between contacts and symptoms was at most very weak, with relatively little variation in contacts alongside substantial variation in symptoms, perhaps with the exception of contact between children. Although there were “Christmas peaks” in adult contact rates during the last two seasons studied, the relative increases in respiratory symptoms during those time periods were much larger in magnitude. This study therefore highlights the complex interplay between social behaviours, demographics, and symptom prevalence in the context of the evolving pandemic, and plausibly respiratory infections in general. These findings may help inform public health policy and infection modelling, emphasising the importance of age-stratified approaches and the impact of measures such as school closures on both social behaviours and symptom rates.

## Data availability

De-identified study data are available for access by accredited researchers in the ONS Secure Research Service (SRS) for accredited research purposes under part 5, chapter 5 of the Digital Economy Act 2017. For further information about accreditation, contact or visit the SRS website. Further details about the dataset are available at: *Office for National Statistics; University of Oxford, released 03 April 2023, ONS SRS Metadata Catalogue, dataset, COVID-19 Infection Survey - UK*, https://doi.org/10.57906/r47r-1735

## Declarations

The views expressed are those of the authors and not necessarily those of the National Health Service, NIHR, Department of Health, or UKHSA. This work contains statistical data from ONS which is Crown Copyright. This work was undertaken in the Office for National Statistics Secure Research Service using data from ONS and other owners and does not imply the endorsement of the ONS or other data owners. The use of the ONS statistical data in this work does not imply the endorsement of the ONS in relation to the interpretation or analysis of the statistical data. This work uses research datasets which may not exactly reproduce National Statistics aggregates. The funder/sponsor did not have any role in the design and conduct of the study; collection, management, analysis, and interpretation of the data; preparation, review, or approval of the manuscript; and decision to submit the manuscript for publication. All authors had full access to all data analysis outputs (reports and tables) and take responsibility for their integrity and accuracy. For the purpose of Open Access, the author has applied a CC BY public copyright licence to any Author Accepted Manuscript version arising from this submission.

## Acknowledgements

We wish to thank all the individuals who participated in the COVID-19 Infection Survey.

We are grateful for the support of all the COVID-19 Infection Survey team:

Office for National Statistics: Sir Ian Diamond, Emma Rourke, Ruth Studley, Nick Taylor, Tina Thomas, Fiona Dawe;

Office for National Statistics COVID Infection Survey Analysis and Operations teams in particular Dawid Pienaar, Joy Preece, Sarah Crofts, Lina Lloyd, Michelle Bowen, Daniel Ayoubkhani, Russell Black, Antonio Felton, Megan Crees, Joel Jones, Esther Sutherland;

University of Oxford, Nuffield Department of Medicine: Ann Sarah Walker, Derrick Crook, Philippa C Matthews, Tim Peto, Emma Pritchard, Nicole Stoesser, Karina-Doris Vihta, Jia Wei, Alison Howarth, Kevin K Chau, Lucas Martins Ferreira, Brian D Marsden, Wanwisa Dejnirattisai, Juthathip Mongkolsapaya, Sarah Hoosdally, Richard Cornall, David I Stuart, Gavin Screaton;

University of Oxford, Nuffield Department of Population Health: Koen Pouwels;

University of Oxford, Big Data Institute: David W Eyre, Katrina Lythgoe, David Bonsall, Tanya Golubchik, Helen Fryer;

University of Oxford, Radcliffe Department of Medicine: John Bell;

Oxford University Hospitals NHS Foundation Trust: Stuart Cox, Kevin Paddon, Tim James;

University of Manchester: Thomas House;

UK Health Security Agency: Julie Robotham, Paul Birrell;

Office for Health Improvement and Disparities: John Newton,

IQVIA: Helena Jordan, Tim Sheppard, Graham Athey, Dan Moody, Leigh Curry, Pamela Brereton;

National Biocentre: Ian Jarvis, Anna Godsmark, George Morris, Bobby Mallick, Phil Eeles;

Glasgow Lighthouse Laboratory: Jodie Hay, Harper VanSteenhouse;

Berkshire and Surrey Pathology Services: Muhammad Ehsaan; Eric Haduli, Hugh Boothe, Reggie Samuel;

Welsh Government: Sean White, Tim Evans, Lisa Bloemberg;

Scottish Government: Katie Allison, Anouska Pandya, Sophie Davis;

Public Health Scotland: David I Conway, Margaret MacLeod, Chris Cunningham.

## Author contributions

The COVID-19 Infection Survey was designed and planned by ASW and KBP, and was conducted by ASW, TEAP, PCM, NS, DWE, and the COVID-19 Infection Survey Team. This specific analysis was originally designed by ASW and ED, but with ongoing input from all authors on results. ED conducted the statistical analysis of the survey data. ED and ASW drafted the manuscript and all authors contributed to interpretation of the data and results and revised the manuscript. All authors approved the final version of the manuscript.

## Ethical statement

The study received ethical approval from the South Central Berkshire B Research Ethics Committee (20/SC/0195).

