## Supplementary methods and figures for "Social behaviours and contact patterns across the 2020/21, 2021/22 and 2022/23 winter seasons in the UK, and associations with symptoms"

Adjustment for survey changes**
The effect of the survey layout change on December 14^th^ 2020 was estimated using negative binomial regression, including assessments from November 1^st^ 2020 to February 1^st^ 2021, the period spanning approximately 1.5 months either side of the database change, with a factor smooth (by survey type) for continuous age in years. Effects were estimated separately for each symptom, and these models also included a tensor product (smooth interaction) between age and calendar time, to account for the highly age-dependent time trends observed for most symptoms during this period, when the Alpha variant was emerging in the UK. A longer time period was not included to limit influence from these large fluctuations on estimates. Smooths were estimated using thin plate splines with k=15 basis functions, and fixed effects for day of the week were also included to control for potential recall bias.

The effect of change in data collection method (which occurred over the second half of July 2022) was similarly estimated using assessments from May 1^st^ 2022 to October 1^st^ 2022 (approximately ±3 months), with a factor smooth (by data collection method) for continuous age in years, and otherwise identical model specifications. **Supplementary Figures S1, S2 and S6** illustrate the predicted effect of the changes in survey delivery from these models across symptoms.

One or two age-specific offsets, obtained through the *difference_smooths* function in the *gratia* package in R, were then included in the main models for each study worker assessment conducted before August 2022, correcting to what would have been estimated if remote data collection had been used throughout the survey. Age-specific point estimates of the offsets were used because long running time made it infeasible to use resampling approaches based on their uncertainty for all models. Sensitivity analysis for cough based on the worst case scenario of the offsets being at the lower and upper 95% confidence interval limits showed the impact was relatively modest compared with the size of the estimates **(Supplementary Figure S7).**

**Supplementary Methods
Supplementary Figure 1. Estimated effect of survey changes on symptom reporting across different ages**

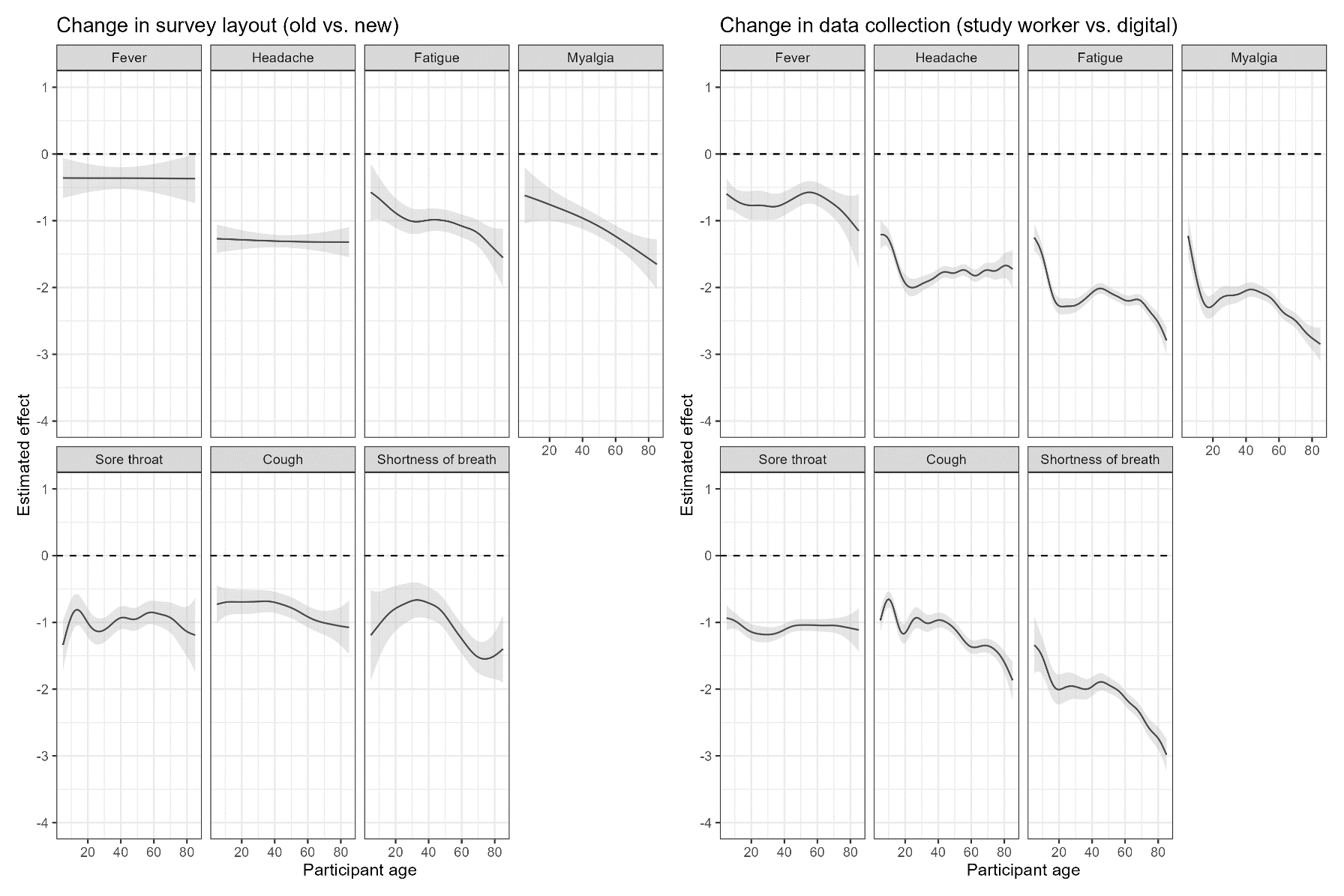

NOTE: Models also included a smooth interaction between age and calendar time, and a fixed effect for day of the week. Effects represent the difference between the old vs. new survey layout, and the difference between study worker and remote data collection (so negative effects reflect lower percentages reporting each symptom before vs after the change). Effects are represented on the log scale (the scale of the negative binomial model coefficient).
**Supplementary Figure 2. Estimated effect of survey changes on contact reporting across different ages**

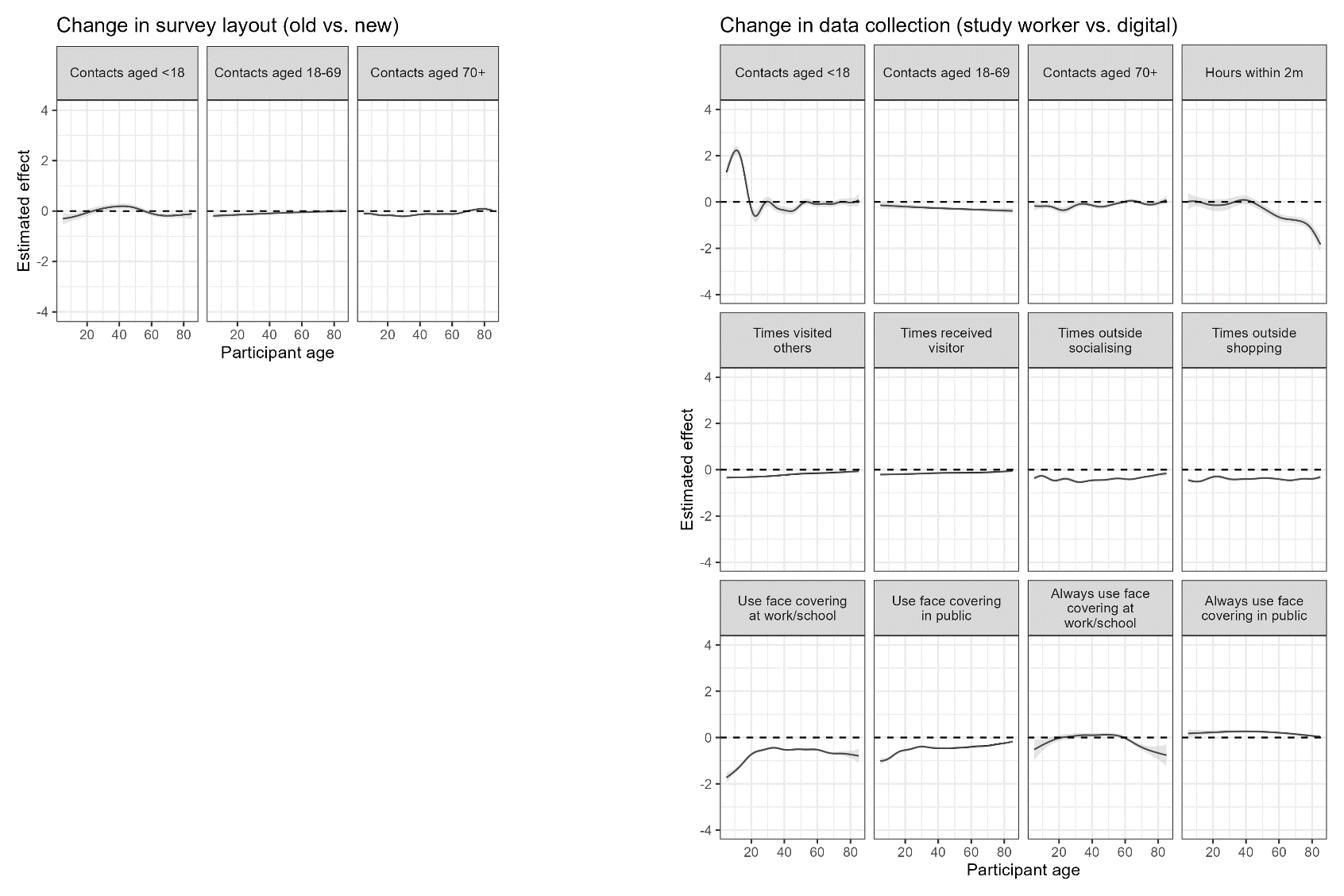

NOTE: Models also included a smooth interaction between age and calendar time, and a fixed effect for day of the week. Effects represent the difference between the old vs. new survey layout, and the difference between study worker and remote data collection (so negative effects reflect a lower number of reported contacts before vs. after the change). Effects are represented on the linear scale, except for the variables related to face covering (log scale).

**Supplementary Figure 3. Trends in remaining contact variables August 2020 – March 2023, by season at specific participant ages**

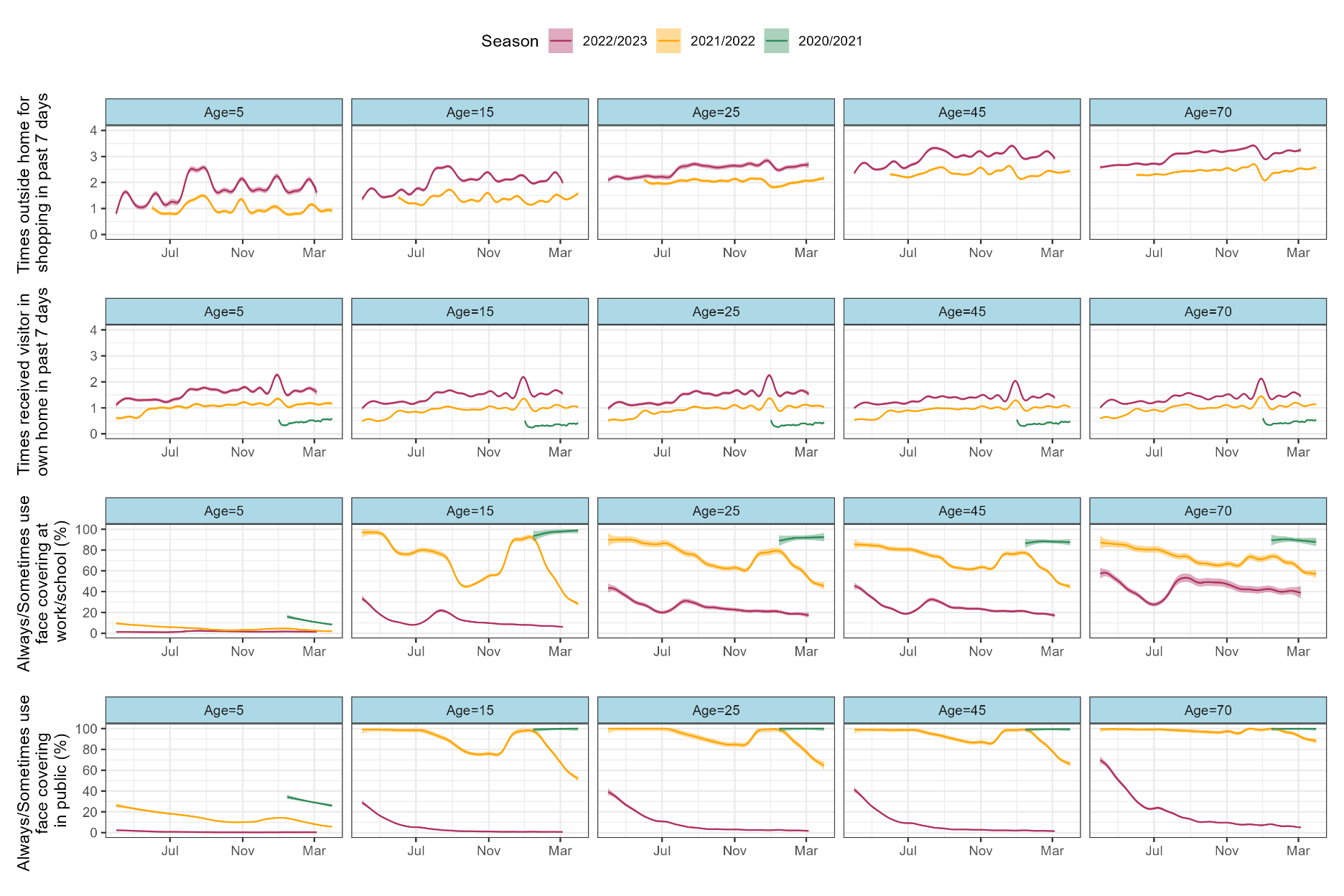

NOTE: Estimates are estimated for the median/mode in-sample levels of the fixed effects (deprivation percentile=63, sex=female, ethnicity=white, country=England, DOW=Wednesday).
**Supplementary Figure 4. Fixed effects for contact variables**

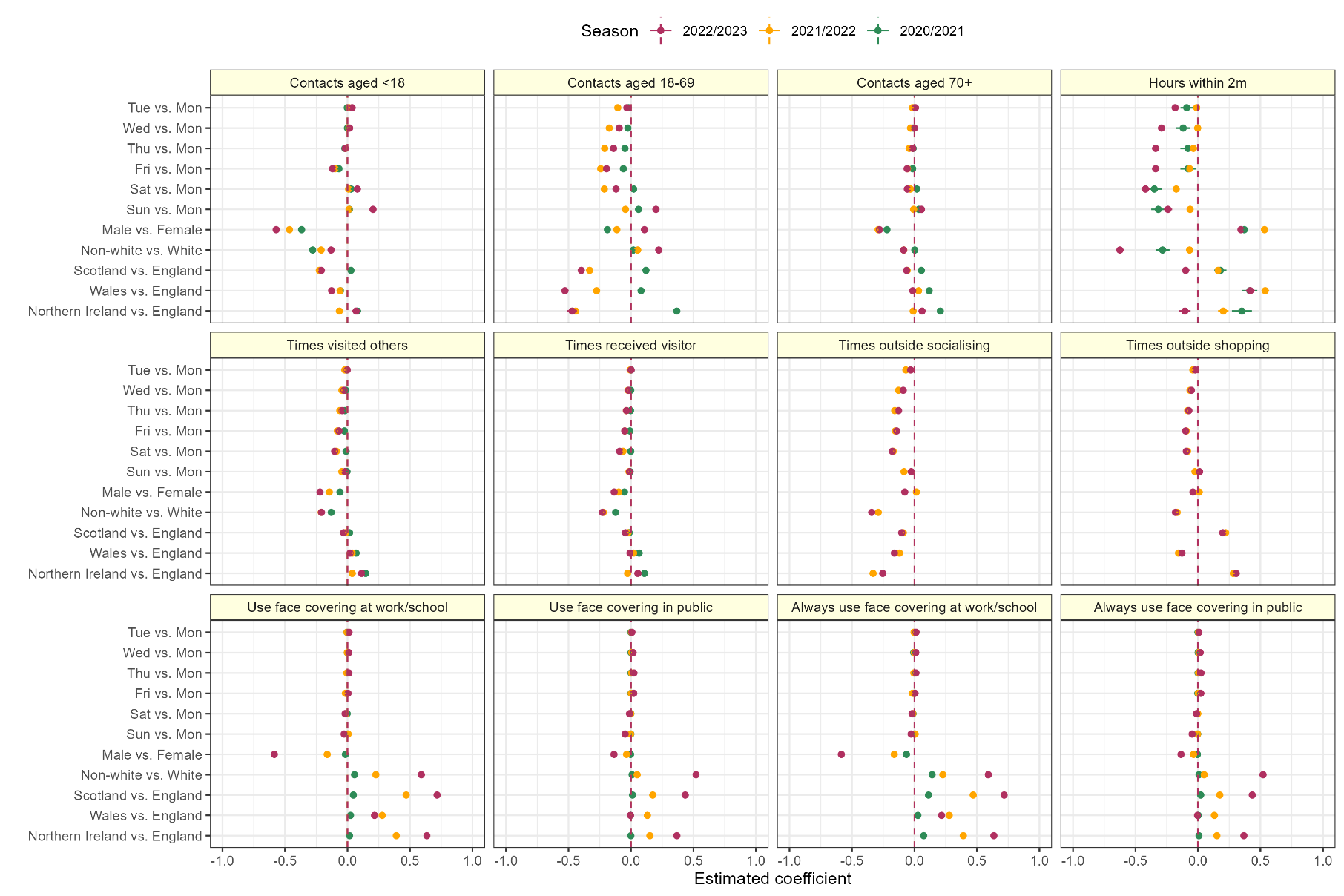

NOTE: Effects are reported on the linear scale for variables on rows 1 and 2, and on the log scale for variables on row 3

**Supplementary Figure 5. Fixed effects for symptom variables**

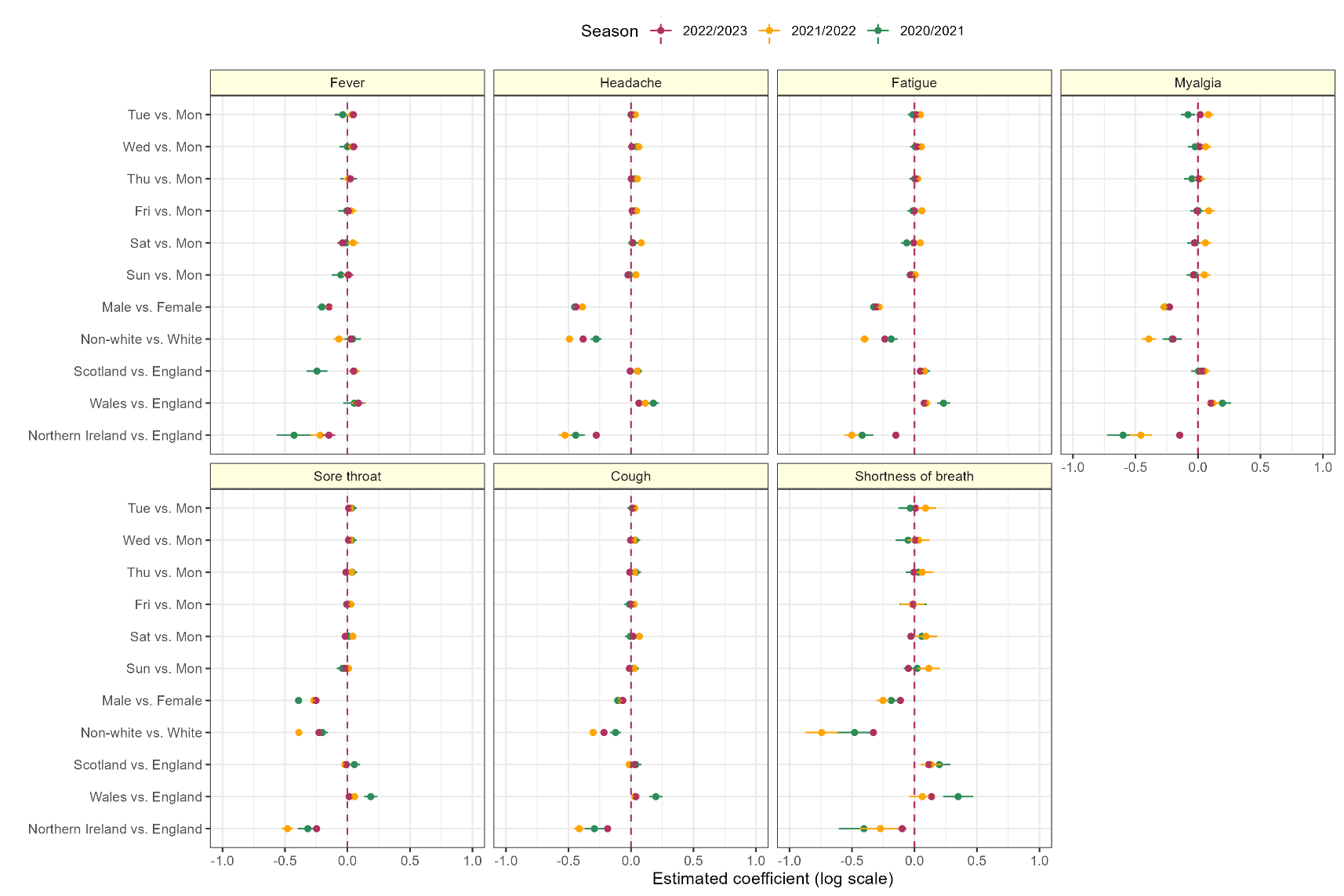

NOTE: Due to computational constraints, models for shortness of breath and myalgia were fit in 1-4 sections per season, effects are illustrated from models containing January 15 in each season (effects were similar across models). **Supplementary Figure 6. Predicted percentages reporting symptoms before and after survey changes, at specific participant ages, without including offsets adjustments**

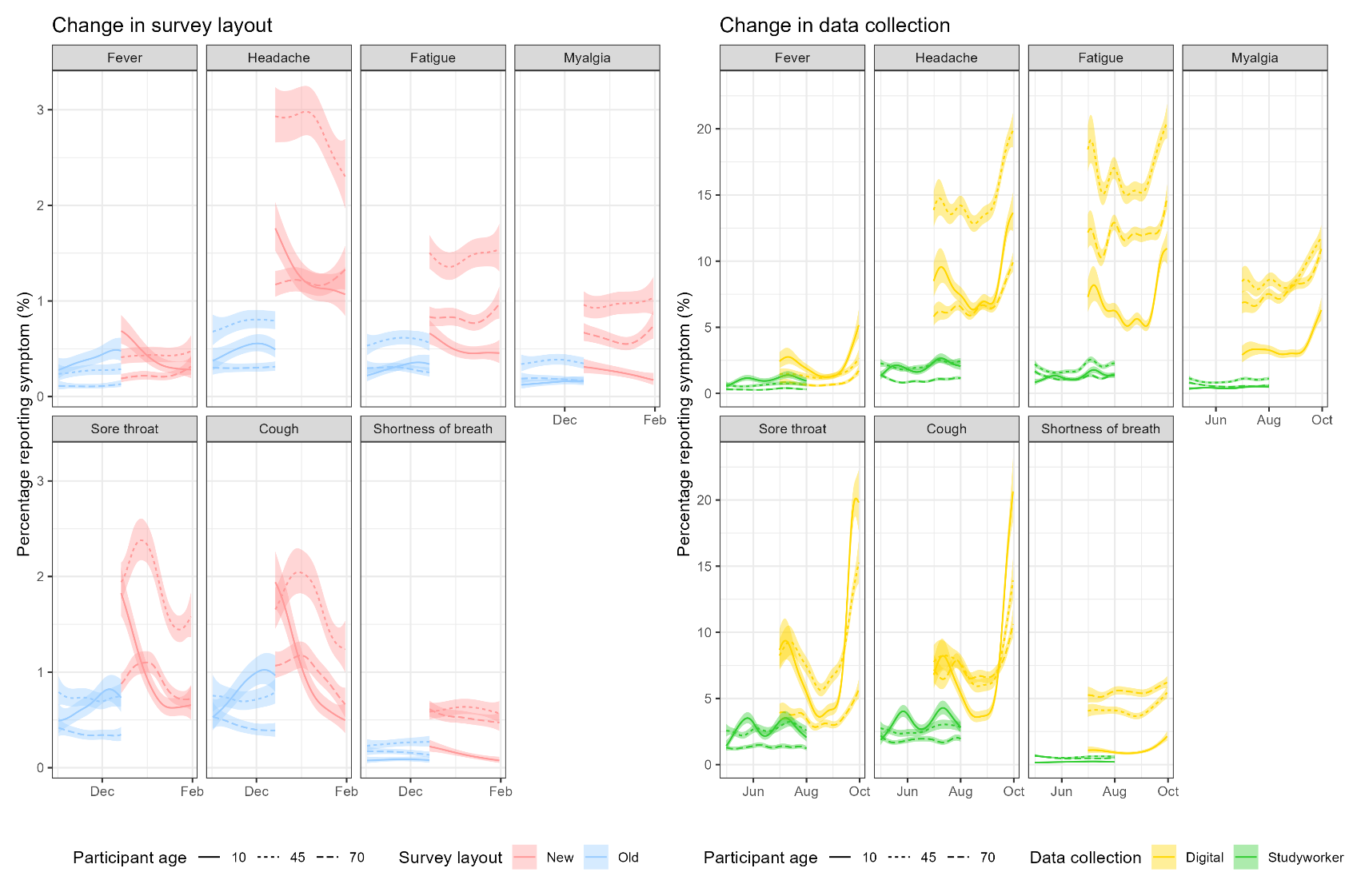

NOTE: Models included a smooth interaction between age and calendar time, and a factor smooth for age by survey layout/collection mode. Fixed effects for day of the week were included, with estimates presented for assessments on Wednesdays.

**Supplementary Figure 7. Sensitivity analysis for survey mode effect adjustment for cough**

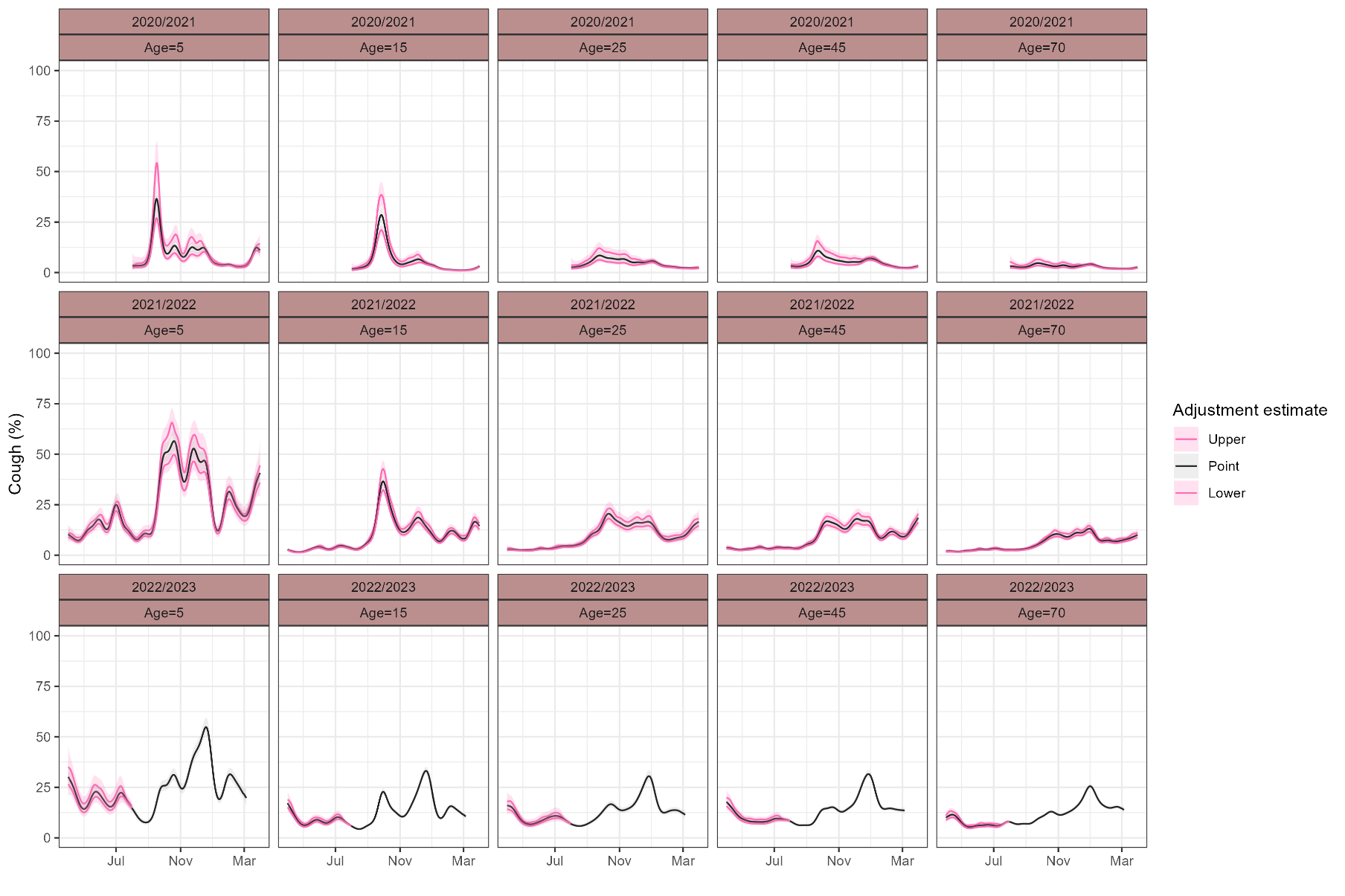

NOTE: “Upper” and “Lower” refer to the 95% confidence intervals for the estimated age-specific effect of survey changes. “Point” refers to the point estimates for the estimated effects, which were used in analysis. Plots are faceted by age and season for ease of interpretation, from August 1^st^ 2022 onwards estimates are identical as all study visits were digital from this date onwards.
